# Whole exome sequencing reveals a wide spectrum of ciliary gene mutations in nonsyndromic biliary atresia

**DOI:** 10.1101/2020.05.05.20091504

**Authors:** Wai-Yee Lam, Man-Ting So, Jacob Shujui Hsu, Patrick Ho-Yu Chung, Diem Ngoc Ngo, Pham Anh Hoa Nguyen, Hannah M. Mitchison, Dagan Jenkins, Christopher O’Callaghan, Pak-Chung Sham, Maria-Mercè Garcia-Barceló, Vincent Chi-Hang Lui, Clara Sze-Man Tang, Paul Kwong-Hang Tam

**Author notes:** **CORRESPONDING AUTHOR**: Paul KH Tam, Division of Paediatric Surgery, Department of Surgery, The University of Hong Kong, Queen Mary Hospital, 102 Pokfulam Road, Hong Kong.

## Abstract

Biliary atresia (BA) is the most common obstructive cholangiopathy in neonates, often progressing to end-stage cirrhosis. BA pathogenesis is believed to be multifactorial, but the genetic contribution remains poorly defined. We conducted exome sequencing on 89 nonsyndromic BA trios. In 31.5% of the patients, rare and deleterious *de novo*, homozygous recessive and/or compound heterozygous variants were detected in liver-expressed ciliary genes of diverse ciliary functions. Enrichment of deleterious mutations in liver-expressed ciliary geneset was significant compared to 148 control trios (OR 2.58, 95% CI 1.15-6.07). *KIF3B, PCNT* and *TTC17* are essential for ciliogenesis. Reduced ciliary proteins expression were detected in the BA livers with *KIF3B* and *TTC17* mutations. CRISPR/Cas9-engineered zebrafish knockouts of *KIF3B, PCNT* and *TTC17* displayed reduced biliary flow. Our findings support a larger genetic contribution to nonsyndromic BA risk than expected. Ciliary gene mutations leading to cholangiocyte cilia malformation and dysfunction could be a key biological mechanism in BA pathogenesis.

## INTRODUCTION

Biliary Atresia (BA) is a major cause of neonatal cholestasis, characterized by progressive fibrosclerosing and inflammatory obliteration of the biliary system during the first few weeks of life. The only current treatment is the Kasai portoenterostomy to restore bile flow. Yet a high proportion of patients, even when bile flow is reestablished, still develop progressive inflammation and sclerosis in the intrahepatic biliary tree, leading to secondary liver cirrhosis. For these patients and those with failed portoenterostomy, liver transplantation is the only treatment option. In neonates, BA is the most common indication for liver transplantation.

BA is rare and varies widely in incidence among populations, being most common in East Asia (1 in 5,000 live births in Asians, compared to 1 in 18,000 live births in Caucasians) (1). BA can be classified into two major groups. The syndromic form, found in 5-20% of BA cases, is characterized by the coexistence of other major congenital anomalies, most commonly laterality defects and splenic malformation. This could be attributed to defective morphogenesis of the bile duct during embryonic development and chromosomal anomalies are a possible genetic origin (2, 3). The nonsyndromic form of BA accounts for 80% of cases, often presenting with later onset of pale stools and jaundice compared to the syndromic form. Nonsyndromic BA is thought to be caused by a combination of environmental insults and genetic predisposition (4). It is hypothesized that a still unknown exogenous factor, possibly viral infection (e.g. cytomegalovirus) or toxins (e.g. biliatresone), meets the innate immune system of a genetically predisposed individual and thereby induces an uncontrollable and potentially self-limiting immune response, which manifests as liver fibrosis and atresia of the bile ducts (5, 6). We conducted the first genome-wide association studies on nonsyndromic BA and identified *ADD3* as a susceptibility gene, showing that common genetic variants could alter disease risk by affecting *ADD3* expression levels in liver (7, 8). Since then, there has been growing evidence that genetic variations contribute to BA risk, with the identification of the additional susceptibility loci *GPC1* (9) and *EFEMP1* (10). Yet, convincing evidence that genetic factors act beyond the role of disease predisposition in the pathogenesis of nonsyndromic BA remains to be found (11).

The occurrence of BA is mostly sporadic, though a limited number of familial cases have been reported previously (12). This could partly be due to the low survival rate into adulthood and the impaired reproductive fitness of BA patients. Thus, if high-penetrance mutations for BA exist, they are most likely to be *de novo*, homozygous recessive or compound heterozygous. In this exome-wide sequencing study, we adopted a family trio design that allowed the identification of exonic *de novo*, homozygous recessive and compound heterozygous variants associated with nonsyndromic BA, with the aim of gaining insights on the molecular or cellular mechanisms underlying its pathogenesis.

## RESULTS

### Most BA cases carry rare and damaging variants in liver expressed genes

Whole exome sequencing (WES) was performed on 91 Southeast Asian nonsyndromic BA family trios, each with one affected patient plus their two unaffected parents. The mean and median sequencing depths were 31.9x and 28x respectively. After quality control, 89 BA family trios remained, as two trios were excluded due to problematic biological relatedness and sample contamination. Thirty-four (38.2%) of the BA subjects were male, and 79 (88.8%) of the subjects had undergone Kasai hepatoportoenterostomy at the average age of 2.2 months (range: 1-7 months).

To identified potentially pathogenic variants, we filtered nonsynonymous variants that were rare, damaging and liver-expressed (RDL), based upon the maximum population minor allele frequency (MAF), liver/biliary tissue and BA organoid gene expression and the deleteriousness prediction of the altered protein. We identified 45 *de novo* mutations, of which 8 were loss-of-function (LOF) variants (6 stopgain and 2 frameshift), in addition to 112 homozygous recessive and 121 compound heterozygous mutations, distributed in 239 genes and 82 BA subjects (92.1%). None of these genes had been linked to BA in previous studies and case reports (11). *PKHD1*, found to have a biallelic variant in one trio of our cohort, was screened in a previous study of perinatal BA patients, but no pathogenic mutation was found (13). There were 3 genes with multiple variant occurrences (observed in >2 subjects in our cohort). Of these, all were predicted as being tolerant to mutation according to the constraint score (missense z-score: *TTN*, -1.10; *PLEC*, -2.57; *USH2A*, -2.47), and could thus be expected to have a larger number of missense mutations observed than predicted by the usual gene mutation rate.

### Liver-expressed genes with rare and damaging variants in BA are enriched for cilium functional annotation

Gene set functional enrichment analysis allows for systematic identification of classes of genes according to biological or molecular function associated with the phenotype of interest, that would not otherwise be detectable on the individual gene level. Within the set of 239 genes with RDL variants, seven gene ontology (GO) biological processes were overrepresented, namely cytoskeleton organization, cilium organization, microtubule cytoskeleton organization, microtubule-based process, cilium assembly, spindle organization, and organelle assembly (Table 1). These functional processes are related and can be regarded as potentially hierarchical in nature. For instance, microtubules are a core component of the cilium axenome for cilia assembly, though they can also localize and function outside the cilia compartment, additionally they are involved in spindle organization during mitosis. The cytoskeleton broadly includes microtubules, microfilaments and intermediate filaments, and the microtubule-based cilium is essential in regulation of ciliogenesis.

**Table 1.** Results of gene set enrichment analysis on 239 genes with RDL variants. List of overrepresented GO biological process and molecular function with *p*-value <0.05 after multiple testing correction using g:Prfoiler’s default mechanism was shown. a, b, c denotes the gene *KIF3B*, *PCNT* and *TTC17* respectively functionally annotated with the enriched GO term.

| Term description | GO term ID | Gene count | $P$ |
| --- | --- | --- | --- |
| **GO biological process** |  |  |  |
| Cytoskeleton organization <sup>a,b,c</sup> | GO:0007010 | 39 | $1.09 \times 10^{-3}$ |
| Cilium organization <sup>a,b,c</sup> | GO:0044782 | 18 | $3.22 \times 10^{-2}$ |
| Microtubule cytoskeleton organization <sup>a,b</sup> | GO:0000226 | 22 | $4.23 \times 10^{-3}$ |
| Microtubule-based process <sup>a,b</sup> | GO:0007017 | 26 | $5.84 \times 10^{-3}$ |
| Cilium assembly <sup>a,b</sup> | GO:0060271 | 17 | $7.24 \times 10^{-2}$ |
| Spindle organization <sup>a,b</sup> | GO:0007051 | 11 | $2.08 \times 10^{-2}$ |
| Organelle assembly <sup>a,b,c</sup> | GO:0060271 | 26 | $3.38 \times 10^{-2}$ |
| **GO molecular Function** |  |  |  |
| Cytoskeletal protein binding <sup>a</sup> | GO:0008092 | 34 | $4.09 \times 10^{-5}$ |
| Cell adhesion molecule binding | GO:0050839 | 20 | $2.82 \times 10^{-3}$ |
| Protein-containing complex binding <sup>a</sup> | GO:0044877 | 31 | $1.68 \times 10^{-2}$ |
| Ankyrin binding | GO:0030506 | 4 | $4.50 \times 10^{-2}$ |

Some of our inputted genes were involved in more than one of these enriched GO functional terms. A total of 49 unique RDL genes were annotated with these seven terms, among which 31 genes (63.3%) were annotated with more than one term, with 18 (58.1%) of the 31 also classed as ciliary genes based on our ciliary gene set. At the extreme, 12 genes were annotated with at least six of the seven enriched terms, and all of them were cilia genes. For GO molecular function, the most significantly enriched term was cytoskeletal protein binding, in line with the biological process findings. Analysis using the KEGG database did not result in any enrichment. Hypergeometric test did not find overrepresentation of ciliary genes in liver-expressed genes in general (*P*=1.0), so that the GO enrichment is specific to genes identified to contain *de novo*, homozygous recessive, or compound heterozygous mutations in BA.

### Ciliary mutation and excess gene set burden

Based on our findings on the overrepresented processes in the exome-wide sequencing data, we hypothesized that the cilium is the key organelle affected by the genetic mutations observed in BA subjects. This led us to further explore and test whether cilia-related processes or components are involved in the pathogenesis of BA. First, we investigated whether BA probands carried an excess burden of mutations in ciliary genes compared to the general population.

The proportion of RDL variants in genes matching the ciliary gene list is summarized in Table 2. As many as 37.5% of protein truncating *de novo* mutations, which were yet to undergo natural selection and can be considered potentially the most damaging, were in ciliary genes (*KIF3B*, *PCNT, SPEF2*) and all were stopgain mutations (Supplementary Table S1). The *KIF3B* and *SPEF2* stopgain mutations identified in the same subject (BA634C) were ultra-rare, not found in the GnomAD, ExAC and 1000 Genomes databases, which together contain >140,000 samples. The case with a *de novo* stopgain mutation in *PCNT* (VBA177C) also harboured an ultra-rare *de novo* missense mutation that was 52bp apart within the same exon on the same parental chromosome. Among the 36 RDL *de novo* missense variants, 8.3% were in ciliary genes *(TTC17, CEP131, PCNT)*. In the RDL homozygous recessive and compound heterozygous variants, 8.9% and 13.2% were in cilia-related genes, respectively. Regardless of the inheritance mode and mutation type, overall 11.5% of RDL variants were in 26 ciliary genes. Strikingly, 31.5% of the BA subjects carried at least one RDL variant in ciliary genes. Multiple occurrence of variants was sparse. A homozygous recessive variant in *DNAH8* (c.2915C>T) was found in two BA subjects (BA596C, BA645C). Compound heterozygous variants were found in *PKD1* and *USH2A* in two and three BA cases respectively. In *SYNE2*, there was a subject with homozygous recessive variant, and another with compound heterozygous mutations. The genetic profiles of RDL ciliary variants in the affected subjects are given in Supplementary Table S1.

**Table 2.** Distribution of RDL variants in ciliary genes observed in the BA probands.

| Type of mutation | RDL variant count |  |  |
| --- | --- | --- | --- |
|  | Overall | Ciliary genes |  |
|  | *n* | *n* | % |
| **De novo** | 45 | 6 | 13.3% |
| *Loss-of-function* | 8 | 3 | 37.5% |
| *In-frame* | 1 | - | - |
| *Missense* | 36 | 3 | 8.3% |
| **Homozygous recessive** | 112 | 10 | 8.9% |
| **Compound heterozygous** | 121 | 16 | 13.2% |
| **Overall** | 278 | 32 | 11.5% |

The mutation burden of RDL *de novo* and homozygous recessive variants in ciliary genes in the BA trios compared to population control trios was assessed by ProxECAT, and logistic regression adjusting for gene set specific counts of rare synonymous variants of the subjects. The consistency of results from the two approaches indicates the robustness of the findings despite the sample size. On the set of all protein coding genes, both tests found no enrichment (odds ratio (OR) [95% confidence interval (CI)]=1.12 [0.93-1.35], *P*=0.241, false discovery rate (FDR) adjusted *P* [*P*-adjusted]=0.241; ProxECAT: *P*=0.364, *P*-adjusted=0.484), showing comparability of the case-control data (Table 3). For the liver expressed ciliary gene set, excess enrichment was consistently demonstrated, and the presence of an RDL ciliary variant elevated the BA risk by 2.6-fold (OR [95% CI]=2.58 [1.15-6.07], *P*=0.021, *P*-adjusted=0.034; ProxECAT: *P*=0.024, *P*-adjusted=0.048). The results for non-liver expressed ciliary genes were mixed. The burden was significant in logistic regression but not in ProxECAT (OR [95% CI]=3.36 [1.16-10.70], *P*=0.026, *P*-adjusted=0.034; ProxECAT: *P*=0.484, *P*-adjusted=0.484).

**Table 3.**
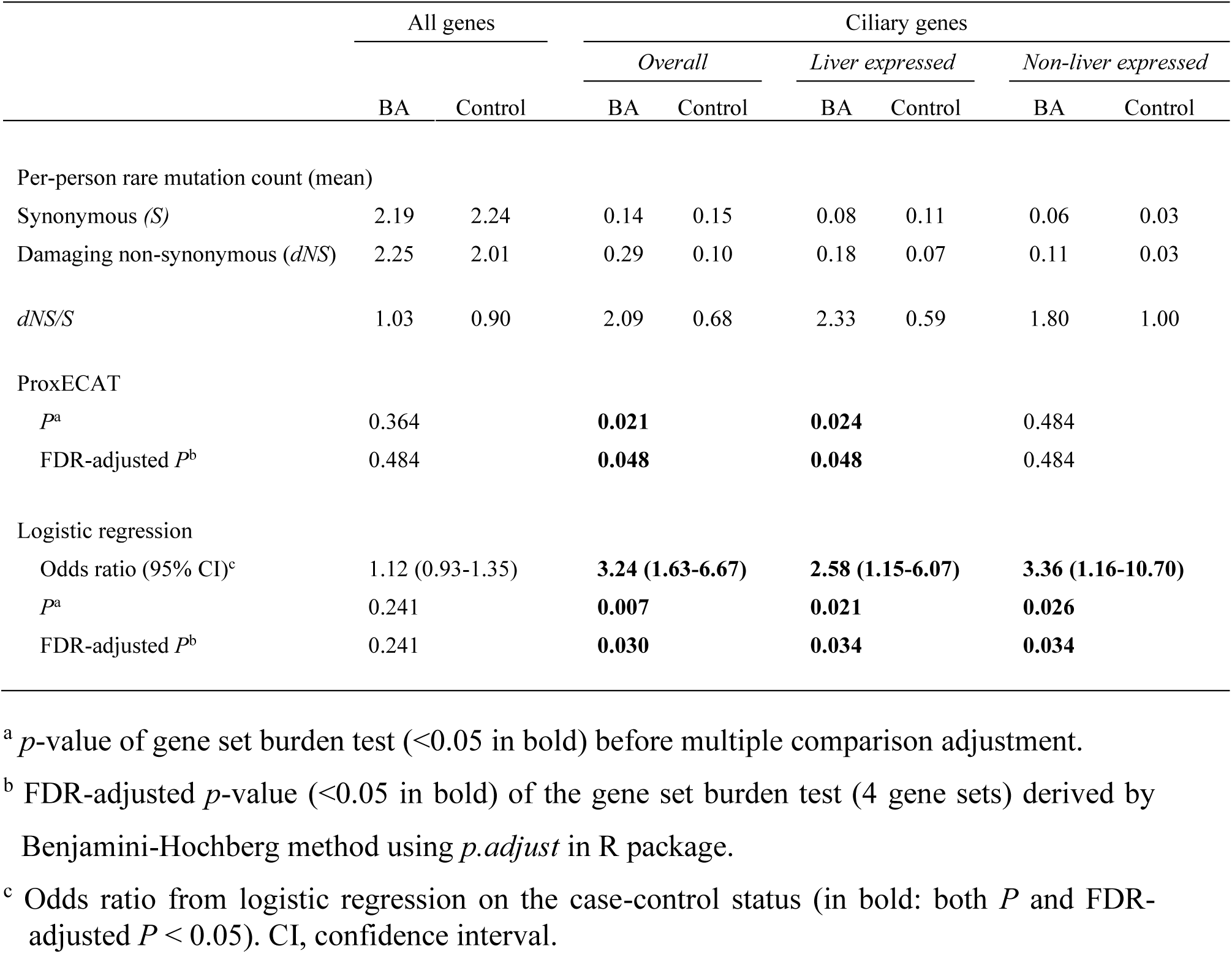
Mutation burden of rare, damaging *de novo* and homozygous recessive variants in 4 gene sets: (a) all protein coding genes, (b) all ciliary genes (n=864), (c) liver expressed ciliary genes (n=586), and (d) non-liver expressed ciliary genes (n=278), in 81 BA trios compared to 148 ethnicity-matched control trios.

This is probably due to sparse variant counts in that gene set (0.03 synonymous/nonsynonymous variant per person is equivalent to 5 counts in the whole control cohort, Table 4) and the difference in statistical power between the two methods. The overall ciliary gene set was dominated by liver expressed genes (67.8%), which could have considerable contribution in the excess burden observed in the overall ciliary gene set in both methods (OR [95% CI]=3.24 [1.63-6.67], *P*=0.007, *P*-adjusted=0.030; ProxECAT: *P*=0.021, *P*-adjusted=0.048).

**Table 4.**
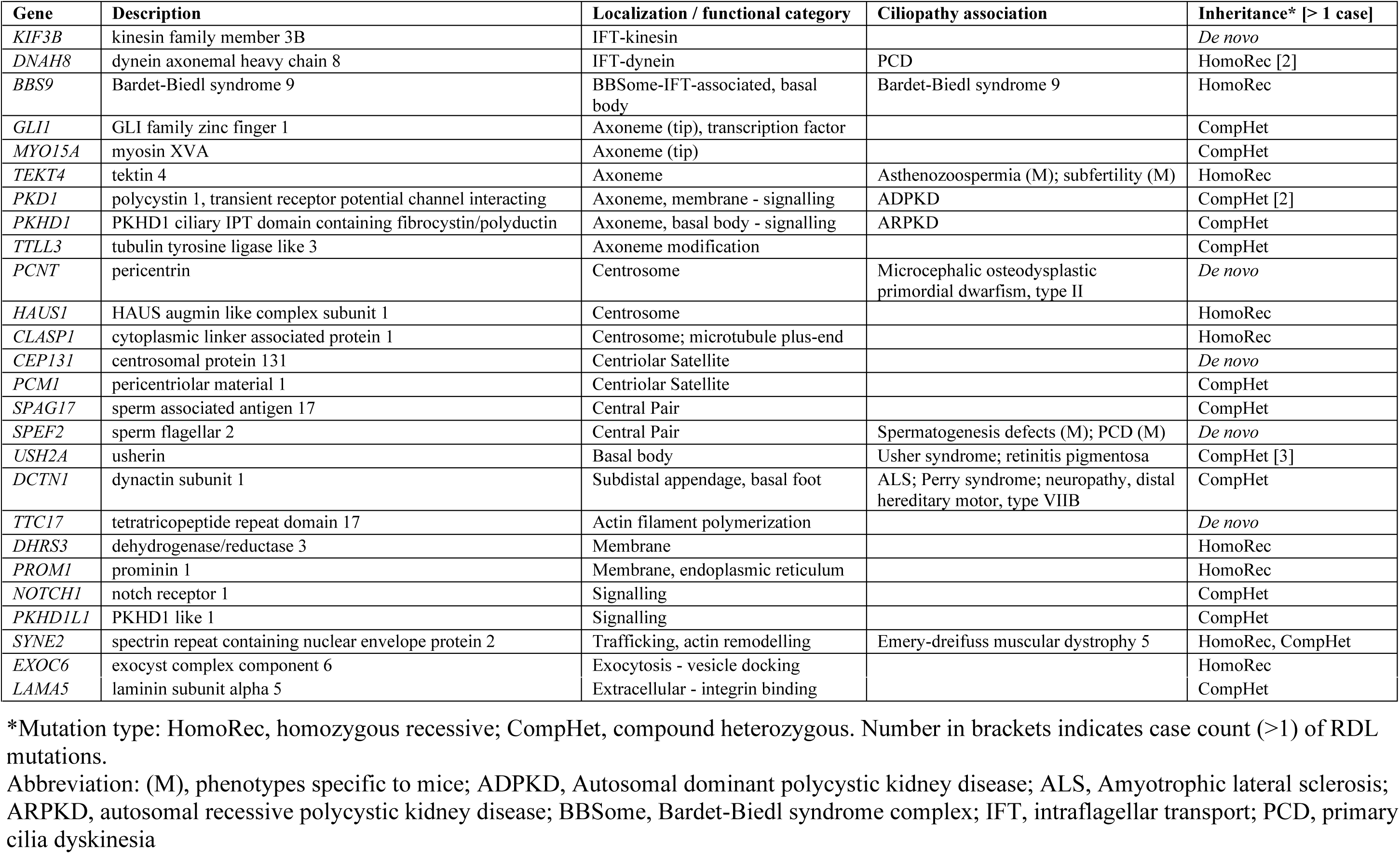
List of 26 ciliary genes with RDL mutations in the BA probands categorized by ciliary localization and functions.

Underlying the excess mutation burden, the 26 ciliary genes identified with RDL variants in the BA subjects represented a diverse spectrum of ciliary functions and localizations (Table 4). Cilia assembly and maintenance depend on intraflagellar transport (IFT) (KIF3B, BBS9) along the microtubules for the trafficking of essential components for cilia formation like DNAH8 and signal transduction proteins. On the axoneme, the GLI1 transcription factor mediates the Hedgehog pathway, while polycystin-1 expressed by *PKD1* forms a ciliary membrane receptor complex for mechanosensation. At the base of the cilia, the basal body, centrosome, centrioles, pericentriolar material and pericentriolar satellites coordinate cilia formation (PCNT, HAUS1, CLASP1, CEP131, PCM1). Several genes encoding proteins localized at the subdistal appendages (DCTN1) and outside the ciliary compartment (TTC17) are essential in pre-ciliogenesis.

We selected three ciliary genes, *KIF3B, PCNT* and *TTC17*, affected by *de novo* mutations in our BA probands, as the representatives of key ciliary localizations and functions in cilia assembly, for the evaluation of the potential of ciliary mutations to cause defective cilia structure and BA phenotypes. KIF3B, the anterograde IFT motor driver, and PCNT at the basal body that forms the pericentriolar material, a nucleation site for microtubules, were both linked to all the seven enriched GO biological processes. TTC17, which is involved in actin organization, localizes outside the ciliary compartment and was linked in two GO overrepresented terms (Table 1).

### Reduced ciliary protein expression in the bile ducts of BA patients

Next we examined if the ciliary mutations correlated with altered ciliary protein expression in the bile duct of the corresponding BA patients. We examined the bile duct expressions of KIF3B and TTC17 of the liver biopsy of the BA subjects carrying *KIF3B* (BA634C) and *TTC17* (BA650C) *de novo* mutations compared to the non-tumor hepatoblastoma tissue of a control subject (Figure 1). In line with the genetic findings, expressions of KIF3B and TTC17 were markedly down-regulated or absent in the bile ducts of the BA subject (BA634C) with *de novo* stopgain in *KIF3B* and *SPEF2*, and the BA subject (BA650C) with *de novo* missense mutation in *TTC17* (Figure 1). Furthermore, reduced expression of cilia marker α-Tubulin was also observed in the bile ducts of the two patients carrying *KIF3B* (BA634C) and *TTC17* (BA650C) *de novo* mutations, suggesting that cilia were not properly formed in these patients’ bile ducts. BA634C also carried a *de novo* stopgain mutation in *SPEF2* (sperm flagellar 2) besides *KIF3B. SPEF2* is known to function in motile cilia, especially for sperm development and tracheal cilia beating, since depletion in *SPEF2* causes male infertility and primary ciliary dyskinesia in mice (14). Therefore the observed down-regulation of α-Tubulin was more likely to be attributed to the *KIF3B* mutation. A liver tissue sample of the Vietnam subject with *PCNT* mutation (VBA177C) was not available, but a previous study reported that PCNT depletion results in loss of primary cilia in human epithelial cells (15).

**Figure 1.**
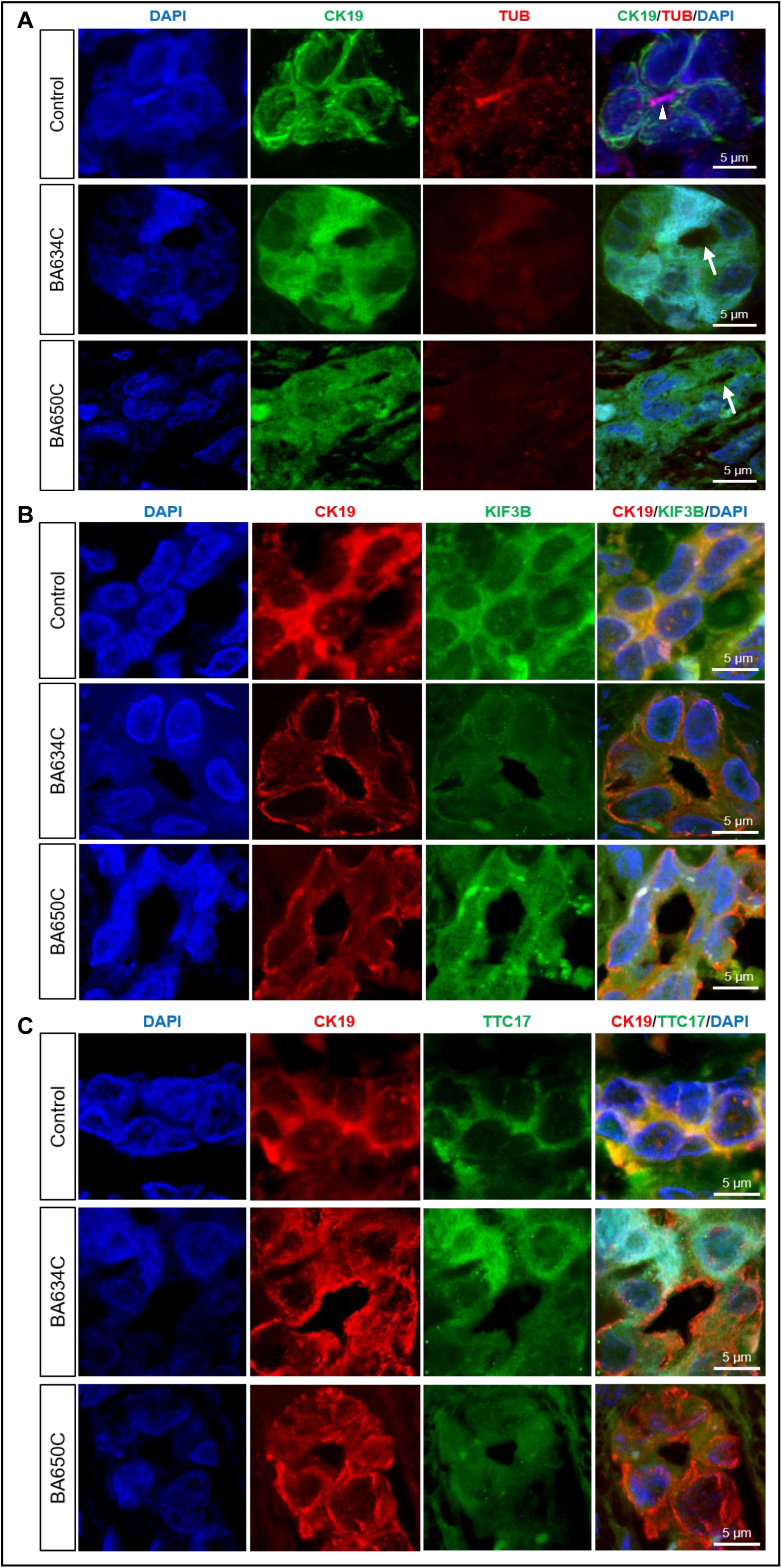
Reduced expression of cilia proteins in the bile ducts of BA liver. Co-immunofluorescence staining for cilia marker α-Tubulin (TUB), KIF3B and TTC17 with CK19 (bile duct marker) were performed on liver sections of the control and BA patients (BA634C and BA650C). (A) α-Tubulin immuno-reactivity (red; arrowhead) was detected at the luminal surface of the bile ducts (CK19 immuno-positive; green) of the control liver. In contrast, expression of α-Tubulin was absent in the bile ducts of BA634C and BA650C (arrows). (B) KIF3B immuno-reactivity (green) was detected at the bile ducts (red) of the control and BA650C, but KIF3B immuno-reactivity was markedly down-regulated in the bile duct of BA634C. (C) TTC17 immuno-reactivity (green) was detected at the bile ducts (red) of the control and BA634C, but TTC17 immuno-reactivity was markedly down-regulated in the bile duct of BA650C. Nuclei were stained with DAPI (blue). All scale bars = 5μm.

### Ciliary gene knockout leads to impaired biliary function in zebrafish

We followed these studies with an assessment of whether ciliary gene depletion could cause the BA phenotype in the zebrafish model. Knockout of *PCNT, KIF3B* and *TTC17* by CRISPR/Cas9-mediated genome editing resulted in defective bile flow in fish embryos, as measured by lowered integrated density of N-([6-(2,4-dinitro-phenyl)amino]hexanoyl)-1- palmitoyl-2-BODIPY-FL-pentanoyl-sn-glycerol-3-phosphoethanolamine (PED6) in the gall bladder in the embryo mutant groups compared to the control group (mean ± standard error of the mean [SEM]: *pcnt*, 1.44 ± 0.13; *kif3b*, 1.24 ± 0.08; *ttc17*, 1.89 ± 0.11; wild type, 2.40 ±0.16; *pcnt* and *kif3b, P* <0.0001; *ttc17*, P=0.017) (Figure 2). This demonstrated that disrupted function of *PCNT, KIF3B* and *TTC17* leads to impaired bile blow in zebrafish model. Introduction of insertion and deletion (INDEL) in *pcnt, kif3b* and *ttc17* genes, and reduced expression of *pcnt, kif3b* and *ttc17* in injected embryo were confirmed by T7 endonuclease I assay and semi-quantitative RT-PCR analysis, respectively (Supplementary Figure S1).

**Figure 2.**
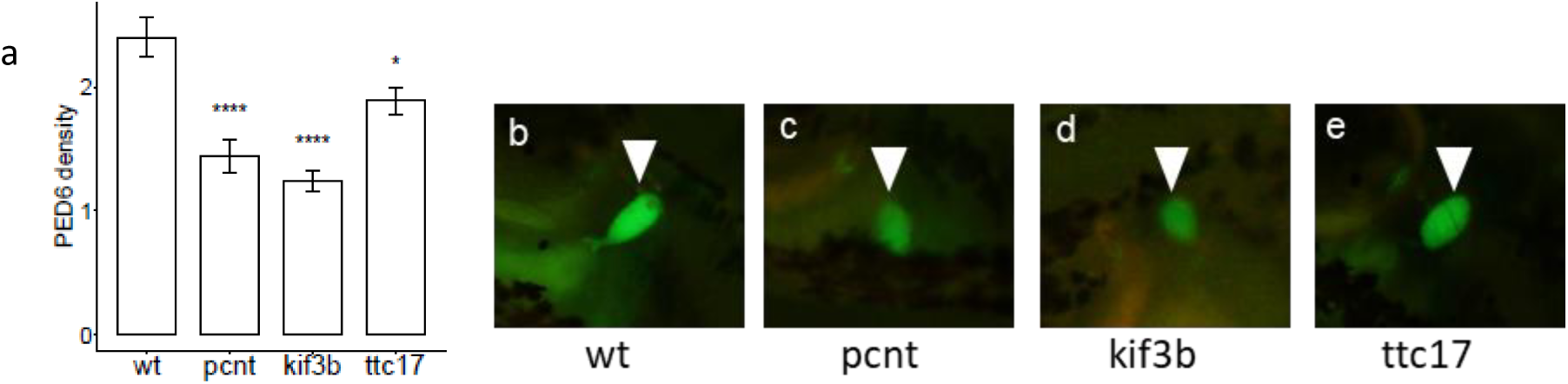
Knockout of *pcnt, kif3b* and *ttc17* resulted in defective bile flow in mutant zebrafish. (a) *pcnt* (n=51), *kif3b* (n=55) and *ttc17* (n=56) mutant embryo group at 5 days post-fertilization showed significantly lower accumulated PED6 integrated density in gallbladder 1 hour after incubation compared to wild type (wt, n=57). Data expressed as mean ± standard error of the mean (SEM). Results of one-way ANOVA with Tukey’s post-hoc test. ****, *P* <0.0001; *, *P* <0.05. (b-e) Representative epifluorescence images showing the mean PED6 integrated density in gallbladders (arrowhead) of each embryo group. (b) wt embryo with gallbladder PED6 integrated density of 2.4. (c) *pcnt* mutant embryo of integrated density at 1.4. (d) *kif3b* mutant embryo with 1.2 integrated density. (e) *ttc17* mutant embryo with integrated density of 1.9.

## DISCUSSION

BA clinically presents with obstruction, inflammation and fibrosis in the biliary tree and is widely considered as multifactorial in origin, involving poorly defined environmental and genetic factors. In this largest WES study on nonsyndromic BA trios to date, we found an excess burden of rare, deleterious mutations carried by BA patients, in a wide spectrum of liver expressed ciliary genes that play different roles in ciliary function and ciliogenesis. Protein expression of the cilia marker and mutated ciliary gene were reduced in our BA patients with *KIF3B* or *TTC17* mutation and the corresponding zebrafish mutants (including *PCNT)* exhibited impaired biliary function. Abnormal cilia formation was caused by the depletion of these genes in other experimental studies (15-18). Based on these findings, we suggest that genetic factors play a primary causal role in a substantial proportion of nonsyndromic BA cases, rather than being merely susceptibility factors. Our findings also implicate defective ciliary structure and function as one of the underlying mechanisms of BA pathogenesis.

Cholangiocyte primary cilia, which protrude from the apical membrane into the ductal lumen, are the physiological sensor for bile flow, composition and osmolality. Signals from the extracellular environment are detected by ciliary specific and associated receptors, ion channels and sensory signalling molecules localized on the ciliary membrane, and transduced through signalling cascades to regulate intracellular activities for cholangiocyte function (19). The Hedgehog (Hh) pathway is pivotal to organ development including the liver during embryogenesis, and tissue remodelling in response to liver injury. It is dormant in healthy livers, yet upon insults, Hh ligand expression is induced in cholangiocytes and the pathway is reactivated to regulate liver regeneration and repair. The primary cilium is indispensable in the complex Hh signaling pathway both physiologically and biologically, interacting with various components at different points of the signaling cascade. The Hh ligand receptors and transcription factors depend on primary cilia for activation, mediation and suppression, and the core signaling components are localized to cilia, thus requiring IFT for their trafficking to the functional sites for regulatory activities (20).

We propose that ciliary gene mutations lead to the development of BA phenotypes through two interconnected biological mechanisms. It was suggested that defective cholangiocyte cilia structure and function can in itself cause overactivation of the Hh pathway, which promotes dysfunctional tissue repair and leads to hepatic inflammation and fibrogenesis (21). In addition, it is likely that defective cilia would compromise the protective function of immature neonatal cholangiocytes against bile acid insults, leading to chronic liver injury, which can also hyperactivate Hh signalling. Indeed, shorter, misoriented, or less abundant cholangiocyte cilia were commonly observed in several studies of both syndromic and non-syndromic BA patients (22-24). Aberrant or absent cilia, or mutations in IFT proteins were shown to cause Hh loss- or gain-of-function phenotypes in different tissue types (25, 26). In cholangiocytes, excessive Hh activation as seen in chronic liver injury triggers epithelial mesenchymal transition and excessive extracellular matrix deposition, thereby leading to biliary fibrosis (21, 27), a prominent pathological feature of BA. In parallel this stimulates immature cholangiocytes to secrete chemokines through autocrine or paracrine mechanisms, which would further recruit, attract and retain immune cells including macrophages, monocytes, neutrophils, and lymphocytes into the injured liver tissues, giving rise to inflammatory response (28), which also feature in BA patients. Evidence of excessive Hh pathway activity and epithelial mesenchymal transition have previously been observed in the extra and intrahepatic ductular cells of nonsyndromic BA patients (29).

In our BA cohort, we identified variants in ciliary genes that are directly involved in the evolutionary conserved IFT process and/or cilia formation, both being indispensable biological processes for Hh signalling. One of the genes carrying variants associated with BA in this study, *KIF3B*, encodes the anterograde kinesin IFT motor and its inhibition was shown to cause cilia loss and overactivation of downstream Hh transcriptional effectors (18). PCNT promotes cilia assembly (15) and PCM1, a component of the centriolar satellites, is required for an efficient Hh response (30). To initiate ciliogenesis, TTC17 modulates actin polymerization for the migration of the basal body to the apical plasma membrane for docking (16) and DCTN1 is required in subdistal appendage organization and microtubules anchoring to the mother centriole (31). GLI1 is a transcription factor of Hh pathway that needs the presence of primary cilia for maximum activation (32). Our proposed pathogenesis is further supported by the studies on two BA susceptibility genes, *ADD3* and *GPC1*, since depletion of either resulted in overexpression of Hh target genes and biliary anomalies and impaired function in zebrafish, that were rescued by Hh inhibition (9, 33). *PKD1L1*, recently identified as a candidate gene in syndromic BA with splenic malformation (34), is also known to regulate downstream Hh signal transduction by forming a component of the ciliary calcium channel, besides involving in the left-right axis specification that is possibly associated with laterality defects (35).

We suggest that the neonatal extrahepatic bile duct can be viewed as serving as the “environmental” factor in BA that, when aggravated with defective cilia structure and function, could trigger chronic liver injury upon the commencement of feeding after birth. The neonatal extrahepatic bile duct is immature structurally and functionally compared to that of the adult and therefore more susceptible to bile acid injury and fibrosis. The protective features in neonatal cholangiocytes, including apical glycocalyx, cell-cell junctions, and structural proteins like collagen and elastin, are less abundant or underdeveloped, while the presence of fibrosis primed cells is excessive (36). Bile acid is able to inflict biliary injury (37), and that might happen at the time when bile flow reaches the intestine during embryonic days 75-85. Several clinical observations are in line with our suggestions. Serum bilirubin levels shortly after birth were found to be higher in newborns later diagnosed with BA compared to healthy controls, indicating bile duct damage beginning in utero (38). Then at the commencement of milk feeding soon after birth, corresponding changes in bile flow and secretion exacerbate the biliary injury in the neonatal bile duct, matching the clinical features of nonsyndromic BA with jaundice onset and stool colour change, worsening in the first few months of life.

The genetic contributions to cholangiocyte ciliary malformation and ciliary protein dysfunction appear to be heterogeneous, demonstrated by the diverse spectrum of liver expressed ciliary genes identified to carry predicted pathogenic variants in BA cases in this study, with scarce multiple occurrences observed for individual variants or genes. We found almost a third of BA subjects with rare, damaging, liver expressed ciliary gene variants, whilst a previous descriptive histopathological study observed cilia abnormalities in 86% of BA subjects (23). The considerable gap could be attributed to the ciliary gene list we applied is relatively conserved, including only genes from SYSCILIA gold standard and GO databases for a list of confident ciliary genes, and not candidate genes in other databases where stronger evidence of their ciliary nature are yet to be established. It could also be that the genetic variations in the non-coding regions contribute to cilia abnormality. Further genetic studies using whole genome sequencing along with further functional studies to delineate the complex effects that perturbation of cilia structure and function have on the Hh signaling pathway in cholangiocytes in relation to the BA phenotype, are warranted.

To conclude, our findings indicate that genetic factors have a more direct role in nonsyndromic BA pathogenesis than previously thought. The excess burden of mutations in liver expressed ciliary genes, conferring potentially reduced ciliary protein expression affecting cilia assembly in patients’ bile duct, implicate that malformed cilia or reduced ciliary function in the vulnerable neonatal biliary system can lead to the over-activation of the Hh signaling pathway especially when exposed to bile acids, resulting in biliary fibrosis, inflammation, and eventually chronic liver injury. Elucidation of disease mechanisms of BA may help in the development of preventative or/and therapeutic strategies to improve the clinical outcome.

## METHODS

### Subjects

A total of 91 unrelated nonsyndromic BA patients and their unaffected parents from the Southeast Asian population participated in the study, of which 45 trios were recruited from Queen Mary Hospital, Hong Kong and 46 trios from the National Hospital of Pediatrics in Vietnam. BA was diagnosed by hepatobiliary scintigraphy and operative cholangiography. The study protocol was approved by the Institutional Review Board of the University of Hong Kong - Hospital Authority Hong Kong West Cluster (UW 05-282 T/945). Informed consent, or informed parental consent for those under 18 years old, was obtained from all participants.

### Whole exome sequencing and bioinformatics analysis

We performed WES on genomic DNA using the xGen Exome Research Panel v1.0 (xGen Lockdown^®^Probes) (83 BA trios) or TruSeq Exome Enrichment Kit v1.0 (8 BA trios) for exome enrichment, sequenced using the Illumina HiSeq 2000 platform at the Centre for PanorOmic Sciences, University of Hong Kong. Sequence reads were aligned by BWA (39) and processed according to the Genome Analysis Toolkit (GATK) best practice (40) version 3.4 for calling single nucleotide variants (SNVs) and small INDELs. Quality control was performed using PLINK (41) and KGGseq (42) (Supplementary Methods).

*De novo*, homozygous recessive and compound heterozygous variants were identified then annotated in KGGseq with the relevant RefSeq gene features, population allele frequencies in public databases, deleteriousness predictions of the altered protein function, and known disease associations from the OMIM and ClinVar databases. To prioritize variants of BA association potential, we focused on nonsynonymous variants that were: (i) rare, (ii) predicted to be functionally damaging and (iii) located in genes expressed in liver or biliary tissues. Population MAF (Supplementary Methods) thresholds applied to filter for rare variants were: <0.005 for *de novo*, <0.01 for compound heterozygous variants, <0.05 for homozygous recessive variants. Variants with the same genotype observed in healthy parents as well as the BA probands were excluded (Supplementary Methods). Functionally damaging variants included all protein-truncating variants and all missense or inframe coding variants that were predicted by SIFT as “deleterious”, by PolyPhen2 as “probably damaging” or that had a CADD score ≥20. Candidate gene mRNA and protein expression in liver or bile duct tissue was checked using the human tissue expression database in EMBL-EBI Expression Atlas (www.ebi.ac.uk) and our in-house BA liver organoid expression database.

### Functional enrichment analysis

Potential disease pathways and molecular mechanisms underlying BA pathogenesis were identified by functional enrichment testing of the gene set (i.e. all RDL variants) within the GO and KEGG databases, using g:Profiler. Overrepresented functional terms were defined by a *p*-value <0.05 after adjusting using g:Profiler’s default multiple testing correction algorithm which has been developed particularly for complex and structured functional profiling data like GO (43).

### Ciliary gene set

The list of ciliary genes was derived by combining: (i) SYSCILIA gold standard cilia gene list (303 genes) (44) and (ii) genes with the annotation “cilium” in the GO human database, searched for using AmiGO (791 genes) (45). The resulting list contained 864 genes involved in ciliary functions with gene products localized within or outside the ciliary compartment (or both), with some genes also exerting additional extraciliary functions. One-tailed hypergeometric test was performed to evaluate any over-representation of ciliary genes in the set of liver-expressed genes.

### Gene set burden test

We compared the mutation burden of rare, damaging variants between 81 (which passed quality check out of 83) BA trios sequenced by xGen Exome Research Panel v1.0 capture kit and 148 ethnicity-matched control trios with non-hepatic congenital condition (8 BA trios enriched by the TruSeq Exome Kit excluded, to minimize any technical variation in sequencing data). Whole genome sequence data of the control trios (mean sequencing depth 39.1x) were processed using identical procedures and software versions to those of the BA variant calling. Quality metrics for sequencing reads and variants in the exonic regions of both cohorts can be found in Supplementary Table S2. Four gene sets were subjected to burden testing: (a) all protein coding genes exome-wide, (b) all selected ciliary genes in our ciliary gene set (864 genes), (c) liver expressed ciliary genes (586 genes), (d) non-liver expressed ciliary genes (278 genes). Burden tests were implemented using ProxECAT, a method formulated by the ratio of the counts of functional to non-functional variants, where the counts of non-functional variants are used as a proxy for the confounding effect that may arise from technical variations (e.g. sequencing techniques) in the case-control data (46). In each gene set, we tested for the difference in the mutation burden of rare, damaging *de novo* and homozygous recessive variants as previously defined, using the rare, synonymous variants in the corresponding gene set as the non-functional proxy (Supplementary Methods). Unless otherwise specified, all statistical analyses in this study were implemented using the R package.

To assess if the BA phenotype was associated with rare, damaging variants, in each of the four gene sets we carried out logistic regression using Firth’s method on the case-control status. We controlled for any background variation effects by including an individual’s count of rare, synonymous variants in the gene set as covariate.

For multiple testing correction, we used the Benjamini-Hochberg method and obtained the FDR-adjusted *p*-value, using the *p.adjust* function in R. An FDR-adjusted *P*<0.05 over the four gene sets was considered to be study-wide significant.

### Liver co-immunofluorescence staining

For BA patients with ciliary gene mutations of interest *(KIF3B* and *TTC17)*, we examined the expression of ciliary proteins KIF3B and TTC17, and cilia marker acetylated α-Tubulin by co-immunofluorescence staining with CK19 (bile duct marker) of liver biopsy specimens obtained for diagnostic histopathology from the Kasai procedure or liver transplantation. Non-tumor liver biopsy specimens from hepatoblastoma subjects were used as controls. Details of the method and antibodies used can be found in Supplementary Methods and Supplementary Methods Table 1.

### Zebrafish genotype-phenotype correlation

We used zebrafish *(Danio rerio)* as an animal model to investigate the *in vivo* association between candidate ciliary gene mutations and the BA phenotype, by CRISPR/Cas9 gene knockout. Guide RNA (gRNA) sequences were co-injected with recombinant Cas9 protein to one-cell stage zebrafish embryos. Injected embryos were collected for genomic DNA extraction and T7 endonuclease I (T7E1) assay to confirm the presence of INDEL mutation (Supplementary Methods). Bile flow was assessed in 51-57 independently injected embryos per wildtype or gene-knockout test group, by measuring the ability to process PED6 (47), a fluorescent lipid reporter for examination of biliary function. We cultured gRNA-injected and un-injected embryos until 5 days post-fertilization, after which they were released to swim in a 5μM solution of PED6 for 2 hours. During this period, images using an Olympus SZX7 fluorescent microscope were taken at 30 minutes intervals.

We analysed the microscopic images using ImageJ (48). The total green fluorescence in the gall bladder was calculated as integrated density (area × mean fluorescence intensity). Comparison of PED6 uptake between each mutant embryo group and wild type was performed using two-tailed one-way ANOVA with Tukey’s post-hoc test. *P*<0.05 was considered to be statistically significant.

## Data Availability

The datasets generated and/or analysed during the current study are available from the corresponding author on reasonable request.

## Data availability

The data of this study are available within the article and in the Supplementary Information, or from the authors upon request.

## AUTHOR CONTRIBUTIONS

W.Y.L. performed the genetics analyses. W.Y.L. and J.S.H contributed to bioinformatics data processing. M.T.S. prepared the samples, performed Sanger sequencing and functional studies. P.H.Y.C., D.N.N, P.A.H.N. recruited the subjects, collected the samples and clinical data. H.M.M, D.J. and C.O. provided insightful comments and suggestions. W.Y.L. wrote the manuscript. P.C.S., M.M.G.B., V.C.H.L., C.S.M.T. and P.K.H.T. designed the study and supervised the project.

## ACKNOWLEDGMENTS

This work was supported by research grants from the Hong Kong General Research Fund (grant no. 17107314 and 766112 to M.M.G.B., 17105119 to V.C.H.L.) and Li Ka Shing Donation Account - Enhanced New Staff Start-up Packages to P.H.Y.C. We extend our gratitude to all the patients and their families who participated in the study, the physicians who referred and treated the patients, and other members of our laboratories for their valuable contributions over the years.

## POTENTIAL CONFLICT OF INTEREST

The authors have declared that no conflict of interest exists.

## SUPPLEMENTARY METHODS

### Bioinformatics processing and quality control

Raw sequence reads were aligned using the BWA-MEM algorithm by BWA. Aligned reads were processed according to the best practice recommendations of the GATK version 3.4. SNVs and INDELs were called by GATK HaplotypeCaller. Validation of biological relatedness of the family trios and evaluation of population stratification of the subjects were conducted with PLINK, and quality control filtering at the variant and genotype level was performed using KGGseq.

### Population minor allele frequency

The population MAF was defined as the maximum of the MAF among all the available population subgroups in the genome aggregation database (gnomAD), Exome Aggregation Consortium database (ExAC) and 1000 Genomes Project database.

### Functionally damaging compound heterozygous variants

Additional criteria were applied in classifying compound heterozygous variants as predicted damaging. This required that at least one of the two identified nucleotide changes in a gene was damaging according to the prediction criteria listed in the main text, that no healthy parent in the cohort was found to be homozygous for either variant and that neither variant was found as an untransmitted allele in other parents in the cohort.

### Burden analysis

Only SNVs were included in the burden analysis because variants in the non-BA control cohort were not validated while INDEL calling are generally of lower accuracy.

### Co-immunofluorescence staining

Paraffin sections (8 μm in thickness) were mounted onto slides and deparaffinised in xylene, followed by rehydration in a series of ethanol. Antigen retrieval was performed by incubation in 1mM Tris-EDTA buffer (pH 9.0) at 95°C for 10 min. After blocking in PBS-T (PBS with 0.1% Tween-20) supplemented with 1% Bovine Serum Albumin for 1 h at room temperature, sections were incubated overnight at 4°C with appropriate primary antibodies according to Supplementary Methods Table 1. Sections were washed three times with PBS and then incubated with Alexa Fluor® 488-conjugated goat anti-rabbit IgG and Alexa Fluor® 594 goat anti-mouse IgG secondary antibodies (1/300, Invitrogen) for 1 h at 37°C followed by PBS washing. Finally, sections were counterstained with DAPI. Samples were imaged on a laser scanning confocal microscopy (Carl Zeiss LSM 880 with Airyscan) equipped with a 63× oil-immersion objective. Images were processed using ImageJ software.

**Supplementary Methods Table 1.** Primary antibodies used for immunostaining of liver tissues.

|  | **CK19** | **α Tubulin** | **CK19** | **KIF3B** | **TTC17** |
| --- | --- | --- | --- | --- | --- |
| **Company** | Abcam | Abcam | Abcam | Biorbyt | Sigma |
| **Catalog number** | ab52625 | ab24610 | ab7755 | orb184776 | HPA038508 |
| **Host** | Rabbit | Mouse | Mouse | Rabbit | Rabbit |
| **Dilution** | 1/400 | 1/100 | 1/200 | 1/300 | 1/300 |

### Generation of zebrafish embryos with the deletion of ciliary gene

Gene deletion was generated by introduction of INDEL into the *pcnt*; *kif3b* and *ttc17* gene via CRISPR/Cas9 and gRNA procedure. The design and synthesis of gRNA were carried out as described (1). CRISPRscan (http://www.crisprs-can.org/) was used to design gRNA sequence against *pcnt*; *kif3b* and *ttc17*, and designs with high predicted efficiency and low predicted off-target effects were chosen. In brief, the forward and the reverse oligos were annealed and end-filled using Taq polymerase. Then the annealed end-filled DNA template was used to generate gRNA specific for the target gene using MEGAshortscript T7 kit (Invitrogen) following manufacturer’s protocol. DNA template was then removed by incubation with TURBO DNase at 37°C for 15 minutes. 1 to 3 nl of injection mixture (sgRNA: 150 pg/nl; Cas9 protein: 667 pg/nl; 0.05% phenol red in RNase free water) was injected into the cytoplasm of one-cell stage um14Tg[Tg(tp1-MmHbb:EGFP)um14] embryos to generate knockout zebrafish larvae. The injected larvae and un-injected control were cultured to 5 days post-fertilization (dpf) for phenotype checking and imaging. Sequences of gRNA were shown in Supplementary Methods Table 2.

**Supplementary Methods Table 2.**
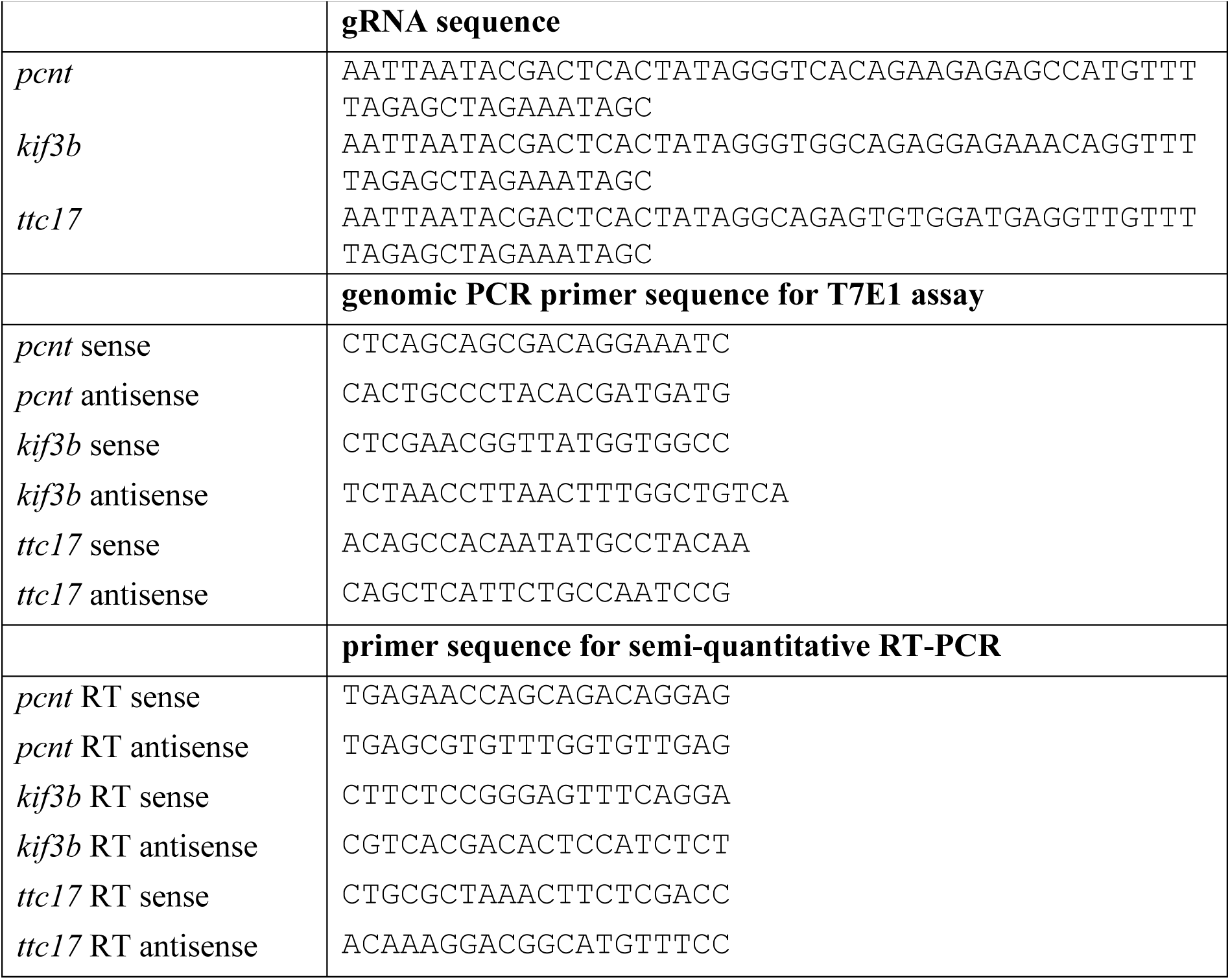
gRNA and primer sequences used in the zebrafish study.

Injected larvae (n=10) were collected for genomic DNA extraction and T7 endonuclease I (T7E1) assay to confirm the presence of INDEL mutation as described previously (2). In brief, the genomic region flanking the sgRNA target site was amplified by PCR. The PCR product was denatured and slowly re-annealed to allow the formation of a heteroduplex. The re-annealed PCR product was digested with T7 endonuclease I (New England Biolab) at 37°C for 45 min and then resolved by 2% agarose gel electrophoresis. Primers for T7E1 assay were tabulated in Supplementary Methods Table 2.

To confirm the successful knockout of *pcnt*; *kif3b* and *ttc17* in knockout embryo that showed defective bile flow, total RNA was extracted from individual 5-dpf embryo injected with the specific gRNA and Cas9 or un-injected (wildtype) using RNeasy Mini Kit (Qiagen) and cDNA were reverse transcribed using PrimeScript RT reagent kit (RR047Q; TakaRa). PCR was performed using a Hot StarTaq (157011236; Qiagen). Expression of a house-keeping gene (beta-actin) was included for comparison. PCR was performed in 25 μl of reaction mixture containing PCR buffer (Roche); MgCl2 (2.5 mM); dNTP (0.2 mM each); forward and reverse primers (0.1 nmole each); 3 μl DNA template and 2.5 units DNA polymerase (QIAGEN; HotStart Taq). The initial denaturation was performed at 95°C for 5 min, PCR reaction was performed for 35 cycles described as follows: After initial 5 min denaturation at 95°C; 30 s denaturation at 95°C, 60 s annealing at specific temperature, 60 s extension at 72°C for each cycle. An additional extension step was carried out for 10 min at 72°C. PCR products were analyzed by electrophoresis in a 1.5 % (w/v) agarose gel and stained by ethidium bromide which was visualized under UV illumination. Primers for RT-PCR analysis of *pcnt; kif3b* and *ttc17* mRNAs were shown in Supplementary Methods Table 2.

---

**Supplementary Table S1.**
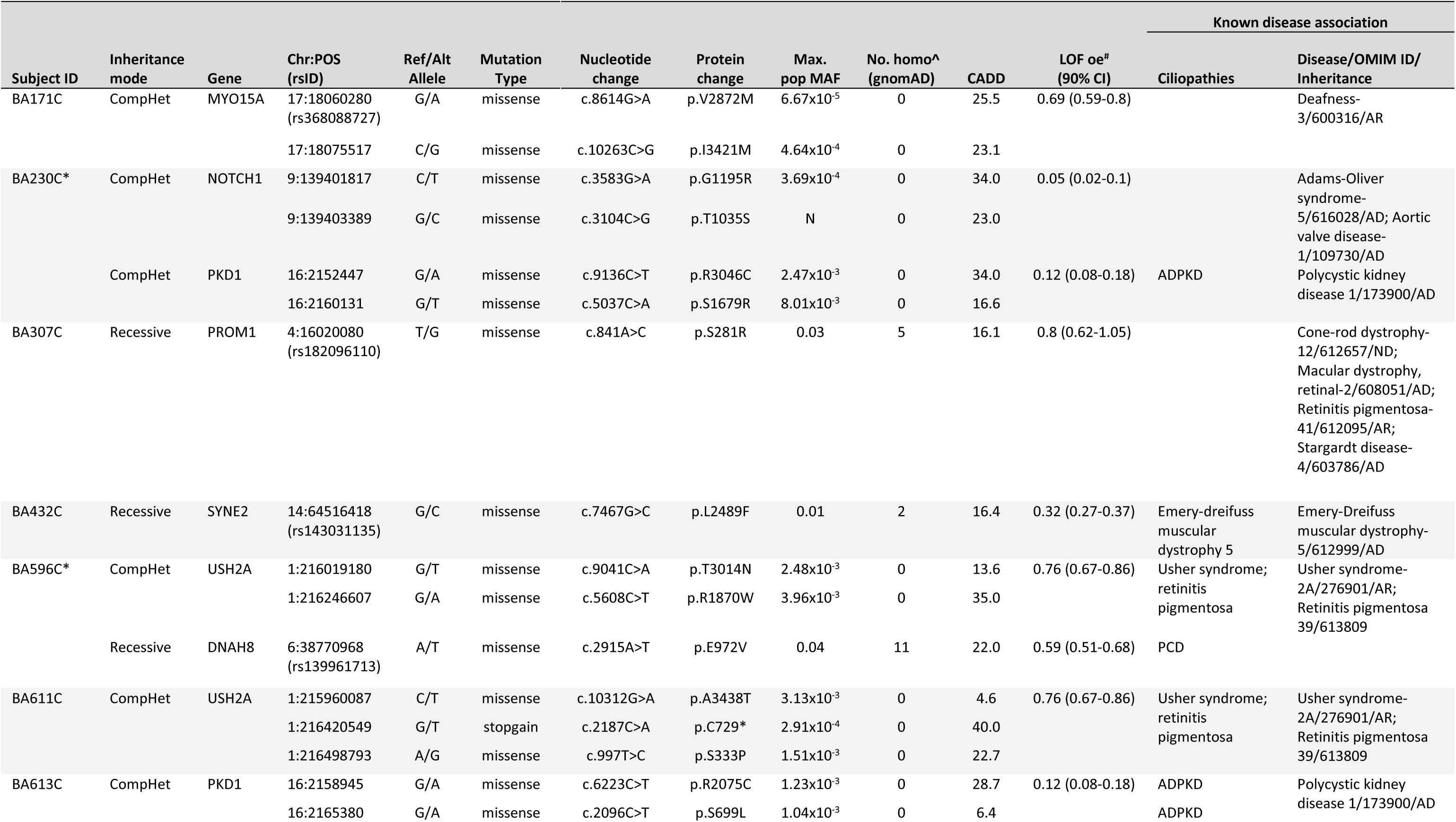
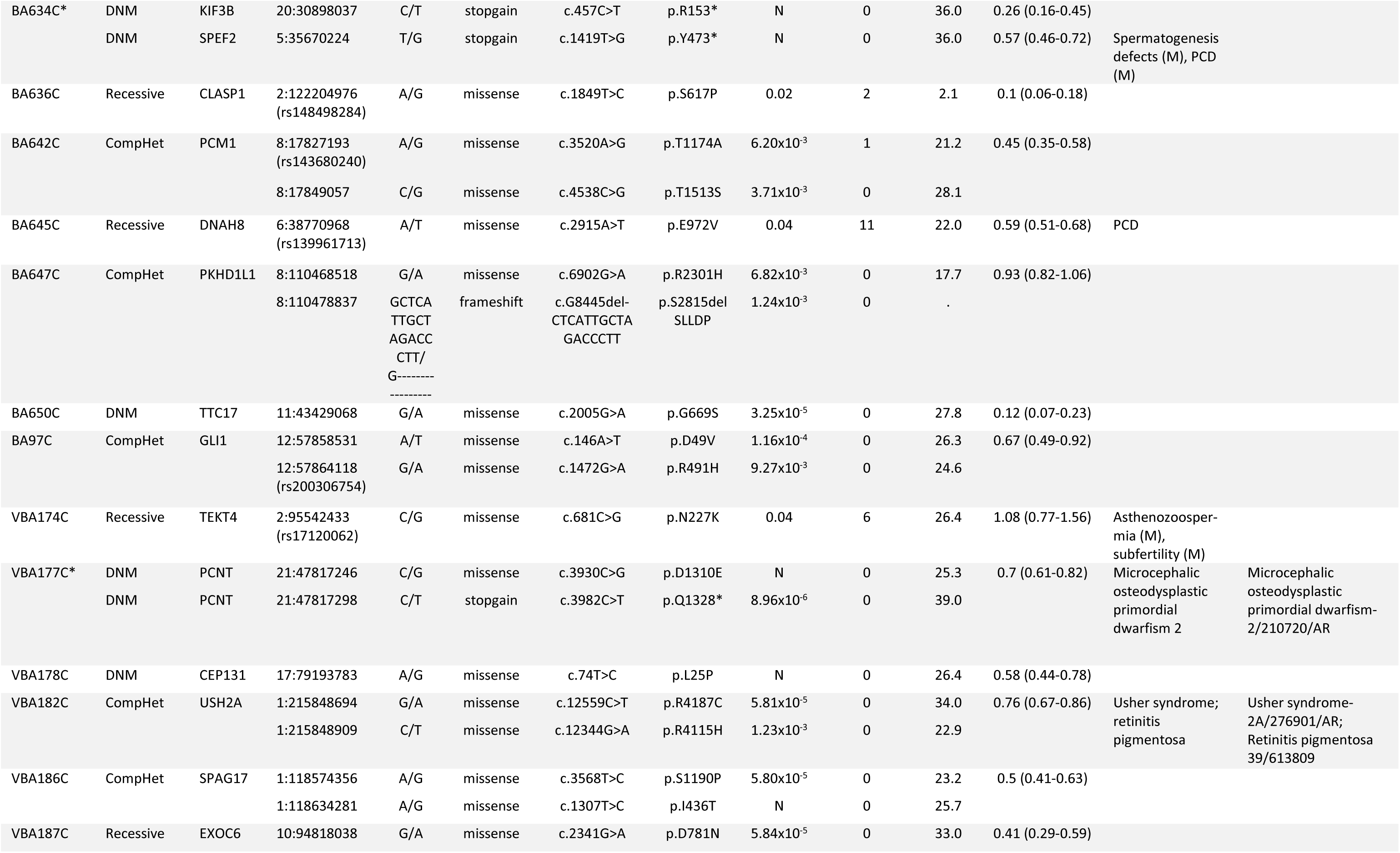
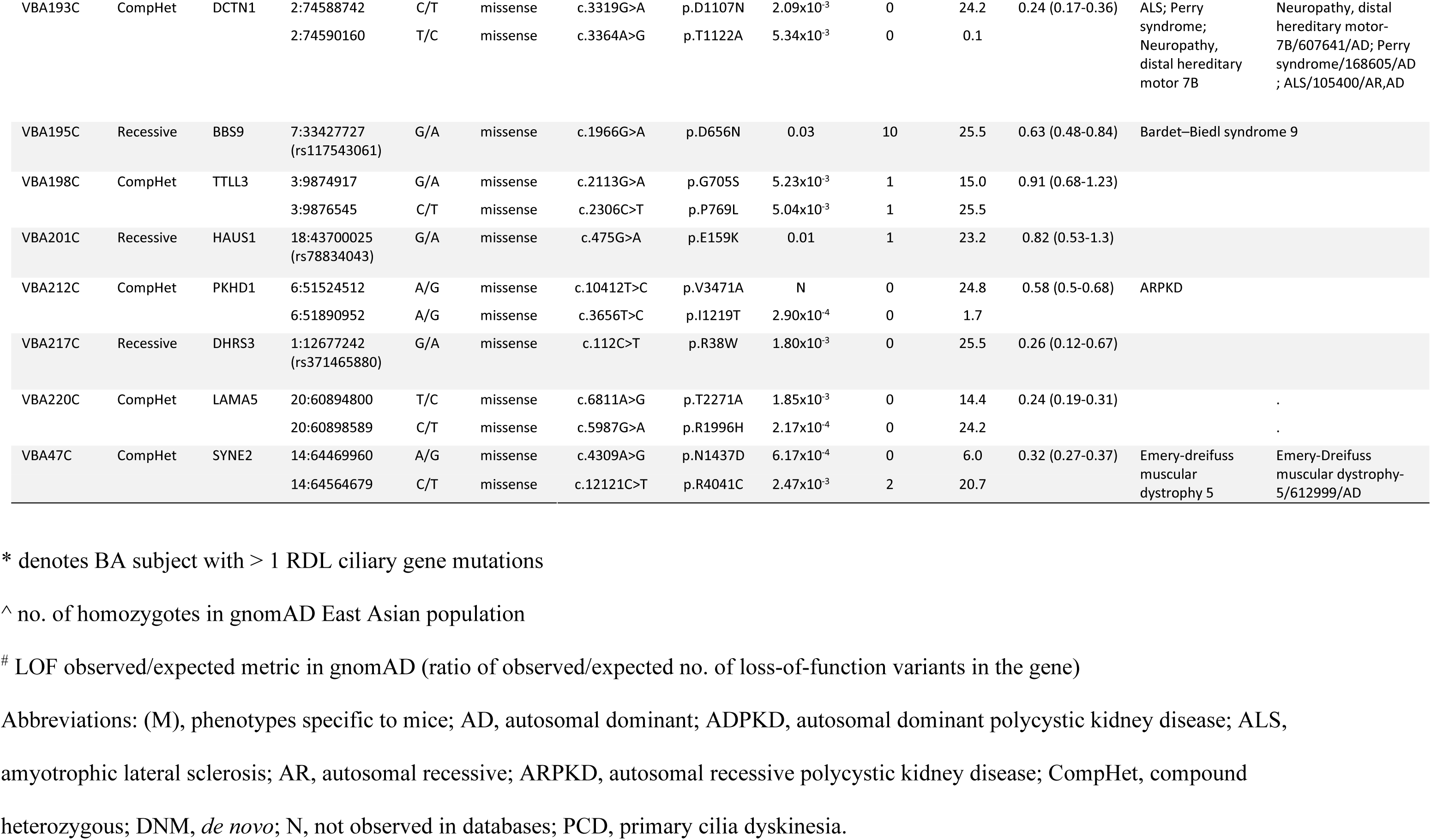
Genetic profile of RDL ciliary variants by subject.

**Supplementary Table S2.**
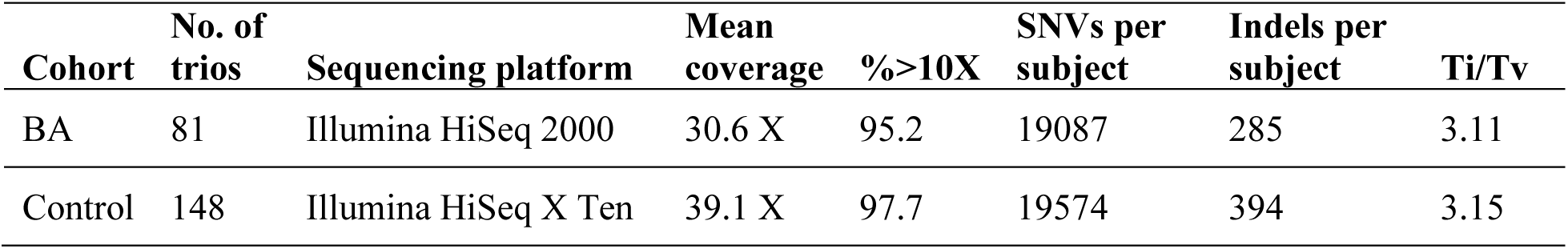
Quality metrics of the sequencing reads and variant calling in the exonic regions for the BA and non-BA control cohorts.

| **Cohort** | **No. of**<br>**trios** | **Sequencing platform** | **Mean**<br>**coverage** | **%>10X** | **SNVs per**<br>**subject** | **Indels per**<br>**subject** | **Ti/Tv** |
| --- | --- | --- | --- | --- | --- | --- | --- |
| BA | 81 | Illumina HiSeq 2000 | 30.6 X | 95.2 | 19087 | 285 | 3.11 |
| Control | 148 | Illumina HiSeq X Ten | 39.1 X | 97.7 | 19574 | 394 | 3.15 |

**Supplementary Figure S1.**
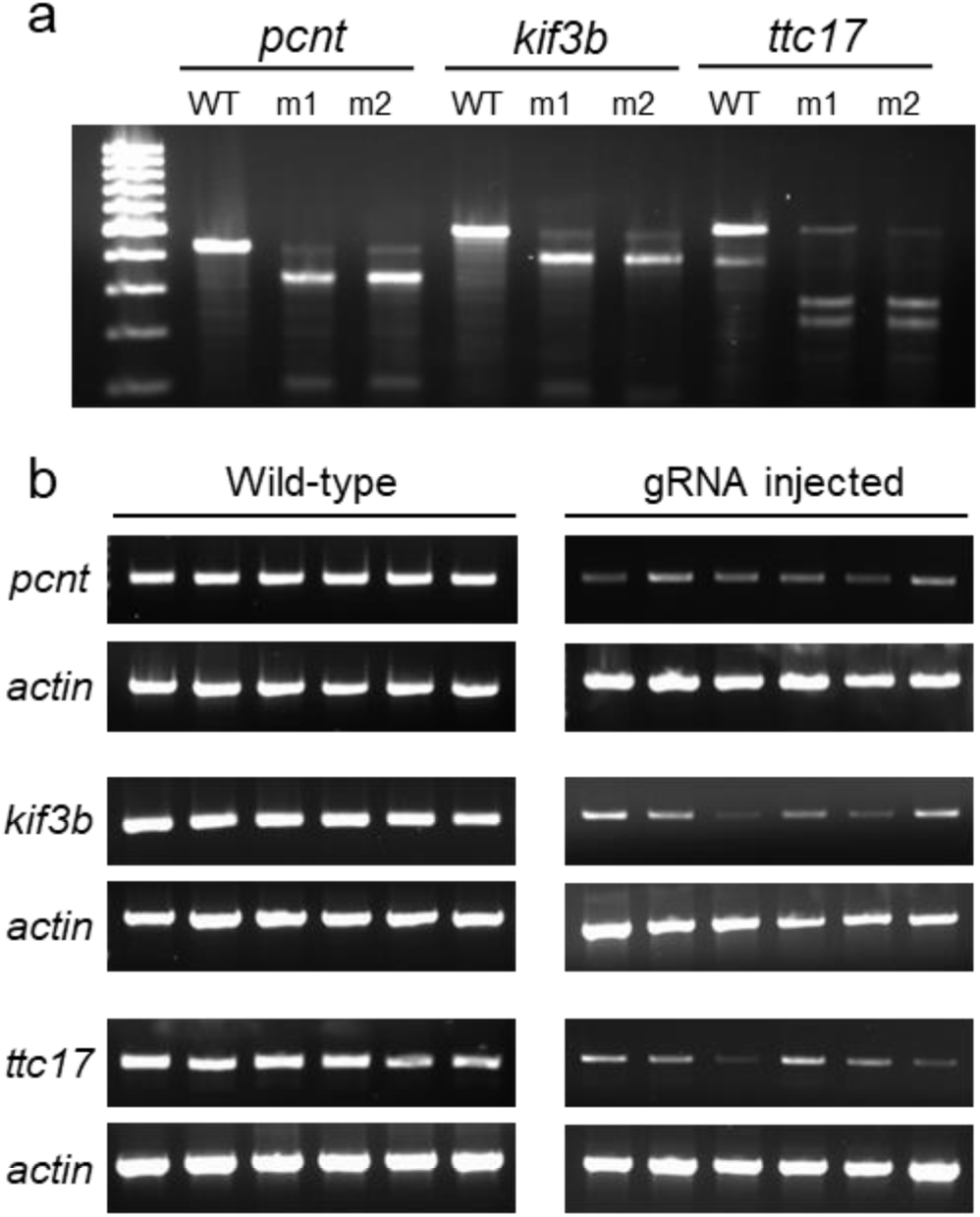
CRISPR/Cas9 gene knockout in zebrafish. Successful creation of INDEL in *pcnt*, *kif3b* and *ttc17* genes were confirmed by T7 endonuclease I assay of genomic PCR products spanning the CRISPR sites from wild-type (WT; n=10) and gRNA injected 5-dpf larvae (ml and m2; n=10) (a). Expression of *pcnt*, *kif3b* and *ttc17* genes in wild-type (WT) and gRNA 5-dpf injected larvae were assayed using semi-quantitative RT-PCR (b). Beta-actin (actin) was used as internal control.

## Notes

### Competing Interest Statement

The authors have declared no competing interest.

## REFERENCES

1. Jimenez-Rivera C, Jolin-Dahel KS, Fortinsky KJ, Gozdyra P, and Benchimol EI. International Incidence and Outcomes of Biliary Atresia. Journal of Pediatric Gastroenterology and Nutrition. 2013;56(4):344–54.

2. Silveira TR, Salzano FM, Howard ER, and Mowat AP. Congenital structural abnormalities in biliary atresia: evidence for etiopathogenic heterogeneity and therapeutic implications. Acta Paediatr Scand. 1991;80(12):1192–9.

3. Tam PKH, Chung PHY, St Peter SD, Gayer CP, Ford HR, Tam GCH, et al. Advances in paediatric gastroenterology. The Lancet. 2017;390(10099):1072–82.

4. Mack CL, and Sokol RJ. Unraveling the Pathogenesis and Etiology of Biliary Atresia. Pediatric Research. 2005;57(7):87–94.

5. Bezerra JA, Wells RG, Mack CL, Karpen SJ, Hoofnagle JH, Doo E, et al. Biliary Atresia: Clinical and Research Challenges for the Twenty-First Century. Hepatology. 2018;68(3):1163–73.

6. Kilgore A, and Mack CL. Update on investigations pertaining to the pathogenesis of biliary atresia. Pediatric Surgery International. 2017;33(12):1233–41.

7. Garcia-Barcelo MM, Yeung MY, Miao XP, Tang CS, Cheng G, So MT, et al. Genomewide association study identifies a susceptibility locus for biliary atresia on 10q24.2. Hum Mol Genet. 2010;19(14):2917–25.

8. Cheng G, Tang CS, Wong EH, Cheng WW, So MT, Miao X, et al. Common genetic variants regulating ADD3 gene expression alter biliary atresia risk. J Hepatol. 2013;59(6):1285–91.

9. Cui S, Leyva-Vega M, Tsai EA, EauClaire SF, Glessner JT, Hakonarson H, et al. Evidence from human and zebrafish that GPC1 is a biliary atresia susceptibility gene. Gastroenterology. 2013;144(5):1107–15.e3.

10. Chen Y, Gilbert MA, Grochowski CM, McEldrew D, Llewellyn J, Waisbourd-Zinman O, et al. A genome-wide association study identifies a susceptibility locus for biliary atresia on 2p16.1 within the gene EFEMP1. PLOS Genetics. 2018;14(8):e1007532.

11. Girard M, and Panasyuk G. Genetics in biliary atresia. Curr Opin Gastroenterol. 2019;35(2):73–81.

12. Smith BM, Laberge JM, Schreiber R, Weber AM, and Blanchard H. Familial biliary atresia in three siblings including twins. Journal of Pediatric Surgery. 1991;26(11):1331–3.

13. Hartley JL, O’Callaghan C, Rossetti S, Consugar M, Ward CJ, Kelly DA, et al. Investigation of primary cilia in the pathogenesis of biliary atresia. Journal of pediatric gastroenterology and nutrition. 2011;52(4):485–8.

14. Sironen A, Kotaja N, Mulhern H, Wyatt TA, Sisson JH, Pavlik JA, et al. Loss of SPEF2 Function in Mice Results in Spermatogenesis Defects and Primary Ciliary Dyskinesia. Biology of Reproduction. 2011;85(4):690–701.

15. Jurczyk A, Gromley A, Redick S, Agustin JS, Witman G, Pazour GJ, et al. Pericentrin forms a complex with intraflagellar transport proteins and polycystin-2 and is required for primary cilia assembly. The Journal of Cell Biology. 2004;166(5):637–43.

16. Bontems F, Fish RJ, Borlat I, Lembo F, Chocu S, Chalmel F, et al. C2orf62 and TTC17 Are Involved in Actin Organization and Ciliogenesis in Zebrafish and Human. PLoS One. 2014;9(1):e86476.

17. Nonaka S, Tanaka Y, Okada Y, Takeda S, Harada A, Kanai Y, et al. Randomization of Left-Right Asymmetry due to Loss of Nodal Cilia Generating Leftward Flow of Extraembryonic Fluid in Mice Lacking KIF3B Motor Protein. Cell. 1998;95(6):829–37.

18. Engelke MF, Waas B, Kearns SE, Suber A, Boss A, Allen BL, et al. Acute Inhibition of Heterotrimeric Kinesin-2 Function Reveals Mechanisms of Intraflagellar Transport in Mammalian Cilia. Curr Biol. 2019;29(7):1137–48.e4.

19. LaRusso NF, and Masyuk TV. The Role of Cilia in the Regulation of Bile Flow. Digestive Diseases. 2011;29(1):6–12.

20. Machado MV, and Diehl AM. Hedgehog signalling in liver pathophysiology. Journal of Hepatology. 2018;68(3):550–62.

21. Omenetti A, and Diehl AM. Hedgehog signaling in cholangiocytes. Curr Opin Gastroenterol. 2011;27(3):268–75.

22. Chu AS, Russo PA, and Wells RG. Cholangiocyte cilia are abnormal in syndromic and non-syndromic biliary atresia. Modern Pathology. 2012;25:751.

23. Frassetto R, Parolini F, Marceddu S, Satta G, Papacciuoli V, Pinna MA, et al. Intrahepatic bile duct primary cilia in biliary atresia. Hepatol Res. 2018;48(8):664–74.

24. Karjoo S, Hand NJ, Loarca L, Russo PA, Friedman JR, and Wells RG. Extrahepatic cholangiocyte cilia are abnormal in biliary atresia. J Pediatr Gastroenterol Nutr. 2013;57(1):96–101.

25. Huangfu D, and Anderson KV. Cilia and Hedgehog responsiveness in the mouse. Proceedings of the National Academy of Sciences of the United States of America. 2005; 102(32): 11325-30.

26. Wong SY, Seol AD, So P-L, Ermilov AN, Bichakjian CK, Epstein EH, et al. Primary cilia can both mediate and suppress Hedgehog pathway-dependent tumorigenesis. Nature Medicine. 2009;15(9):1055–61.

27. Shaheen R, Alsahli S, Ewida N, Alzahrani F, Shamseldin HE, Patel N, et al. Biallelic Mutations in Tetratricopeptide Repeat Domain 26 (Intraflagellar Transport 56) Cause Severe Biliary Ciliopathy in Humans. Hepatology. n/a(n/a).

28. Omenetti A, Syn W-K, Jung Y, Francis H, Porrello A, Witek RP, et al. Repair-related activation of hedgehog signaling promotes cholangiocyte chemokine production. Hepatology. 2009;50(2):518–27.

29. Omenetti A, Bass LM, Anders RA, Clemente MG, Francis H, Guy CD, et al. Hedgehog activity, epithelial-mesenchymal transitions, and biliary dysmorphogenesis in biliary atresia. Hepatology. 2011;53(4):1246–58.

30. Odabasi E, Gul S, Kavakli IH, and Firat-Karalar EN. Centriolar satellites are required for efficient ciliogenesis and ciliary content regulation. EMBO reports. 2019;20(6):e47723.

31. Kodani A, Salomé Sirerol-Piquer M, Seol A, Manuel Garcia-Verdugo J, and Reiter JF. Kif3a interacts with Dynactin subunit p150Glued to organize centriole subdistal appendages. The EMBO Journal. 2013;32(4):597–607.

32. Wang Y, Zeng H, and Liu A. Distinct Activities of Gli1 and Gli2 in the Absence of Ift88 and the Primary Cilia. Journal of Developmental Biology. 2019;7(1):5.

33. Tang V, Cofer ZC, Cui S, Sapp V, Loomes KM, and Matthews RP. Loss of a Candidate Biliary Atresia Susceptibility Gene, add3a, Causes Biliary Developmental Defects in Zebrafish. Journal of pediatric gastroenterology and nutrition. 2016;63(5):524–30.

34. Berauer J-P, Mezina AI, Okou DT, Sabo A, Muzny DM, Gibbs RA, et al. Identification of Polycystic Kidney Disease 1 Like 1 Gene Variants in Children With Biliary Atresia Splenic Malformation Syndrome. Hepatology. 2019;0(0).

35. Delling M, DeCaen PG, Doerner JF, Febvay S, and Clapham DE. Primary cilia are specialized calcium signalling organelles. Nature. 2013;504(7479):311–4.

36. Khandekar G, Llewellyn J, Kriegermeier A, Waisbourd-Zinman O, Johnson N, Du Y, et al. Coordinated development of the mouse extrahepatic bile duct: Implications for neonatal susceptibility to biliary injury. Journal of Hepatology. 2019;[In press].

37. Zhu C, Fuchs CD, Halilbasic E, and Trauner M. Bile acids in regulation of inflammation and immunity: friend or foe? Clin Exp Rheumatol. 2016;34(4 Suppl 98):25–31.

38. Harpavat S, Finegold MJ, and Karpen SJ. Patients With Biliary Atresia Have Elevated Direct/Conjugated Bilirubin Levels Shortly After Birth. Pediatrics. 2011;128(6):e1428-e33.

39. Li H. Aligning sequence reads, clone sequences and assembly contigs with BWAMEM. arXiv:13033997v1 {q-bioGN}.

40. DePristo MA, Banks E, Poplin R, Garimella KV, Maguire JR, Hartl C, et al. A framework for variation discovery and genotyping using next-generation DNA sequencing data. Nat Genet. 2011;43.

41. Purcell S, Neale B, Todd-Brown K, Thomas L, Ferreira MAR, Bender D, et al. PLINK: A Tool Set for Whole-Genome Association and Population-Based Linkage Analyses. The American Journal of Human Genetics. 2007;81(3):559–75.

42. Li MX, Gui HS, Kwan JS, Bao SY, and Sham PC. A comprehensive framework for prioritizing variants in exome sequencing studies of Mendelian diseases. Nucleic Acids Res. 2012;40.

43. Reimand J, Kull M, Peterson H, Hansen J, and Vilo J. g:Profiler—a web-based toolset for functional profiling of gene lists from large-scale experiments. Nucleic Acids Research. 2007;35(suppl_2):W193-W200.

44. van Dam TJ, Wheway G, Slaats GG, Huynen MA, Giles RH, and Group SS. The SYSCILIA gold standard (SCGSv1) of known ciliary components and its applications within a systems biology consortium. Cilia. 2013;2(1):7.

45. Carbon S, Ireland A, Mungall CJ, Shu S, Marshall B, Lewis S, et al. AmiGO: online access to ontology and annotation data. Bioinformatics (Oxford, England). 2009;25(2):288–9.

46. Hendricks AE, Billups SC, Pike HNC, Farooqi IS, Zeggini E, Santorico SA, et al. ProxECAT: Proxy External Controls Association Test. A new case-control gene region association test using allele frequencies from public controls. PLOS Genetics. 2018;14(10):e1007591.

47. Matthews RP, EauClaire SF, Mugnier M, Lorent K, Cui S, Ross MM, et al. DNA hypomethylation causes bile duct defects in zebrafish and is a distinguishing feature of infantile biliary atresia. Hepatology. 2011;53(3):905–14.

48. Schneider CA, Rasband WS, and Eliceiri KW. NIH Image to ImageJ: 25 years of image analysis. Nature Methods. 2012;9:671.

## References

1. Moreno-Mateos MA, Vejnar CE, Beaudoin JD, Fernandez JP, Mis EK, Khokha MK, et al. CRISPRscan: designing highly efficient sgRNAs for CRISPR-Cas9 targeting in vivo. Nat Methods 2015;12(10):982–8.

2. Jao LE, Wente SR, Chen W. Efficient multiplex biallelic zebrafish genome editing using a CRISPR nuclease system. Proc Natl Acad Sci USA 2013;110(34):13904–9.

